# Genetic Liability for Internalizing–Externalizing Traits in Mexican Youth: Moderation by Adversity

**DOI:** 10.64898/2025.11.30.25341187

**Authors:** Zuriel Ceja, I-Tzu Hung, Nathaniel Thomas, Jackson Thorp, Luis M. García-Marín, Carlos S. Cruz-Fuentes, Miguel E. Rentería, Corina Benjet, Jill A. Rabinowitz, Gabriela A. Martinez-Levy

## Abstract

**Objective:** We examined whether common and specific genetic liabilities for internalized (INT) and externalized (EXT) traits predict corresponding disorders in Mexican youth, and whether adversity moderates these associations.

**Methods:** Participants were 1,130 Mexican adolescents (ages 12-17; 54.9% female). Psychiatric diagnoses and adversities were assessed using the World Mental Health Composite International Diagnostic Interview for Adolescents. Genome-wide association summary statistics for 12 INT and EXT traits were analyzed using genomic structural equation modeling (SEM) to derive polygenic scores (PGS) from latent genetic factors. Associations were tested using multinomial logistic regression.

**Results:** The best-fitting genomic SEM bifactor model included one shared internalizing–externalizing factor (INT–EXT-F) and specific factors for internalizing (INT-SF) and externalizing (EXT-SF). Higher INT-EXT-F PGS predicted EXT-only (OR=1.39, 95%CI= 1.16–1.67) and comorbid disorders (OR=1.42, 95% CI= 1.16–1.75). Parental loss moderated associations between EXT-SF PGS and INT-only (OR=1.76, 95% CI= 1.17–2.64) and comorbid disorders (OR=1.79, 95% CI= 1.14–2.81). Cumulative adversity further increased risk, with higher EXT-SF PGS predicting INT (OR= 1.76; 95% CI= 1.17-2.64), EXT (OR= 1.79; 95% CI= (1.14-2.81), and comorbid outcomes (OR= 1.24; 95% CI= 1.04-1.48), and higher INT-EXT-F PGS predicting INT-only disorders (OR= 1.27; 95% CI = 1.07-1.50).

**Conclusion:** These findings emphasize the value of combining genetic and environmental approaches in mental health studies among Mexican youth, broadening the reach and relevance of psychiatric genomic research beyond European populations.

## Introduction

Internalizing (INT) (e.g., anxiety, depression, and/or social withdrawal) and externalizing (EXT) (e.g., conduct disorder, oppositional defiant disorder) disorders are common in adolescents, with prevalences ranging from 8-15% and 10-24%, respectively [1]. INT and EXT disorders often co-occur [2, 3] and are associated with myriad cognitive, socioemotional, and mental health conditions, including lower working memory capacity [4], greater risk of suicidality [5], criminal justice system involvement [6], and worse cognitive and interpersonal functioning [7]. In Mexico specifically, INT and EXT disorders are among the leading factors prompting individuals to seek mental health care [8]. Therefore, investigating the factors that contribute to the emergence of these disorders, particularly in this population, is crucial.

Twin and family studies indicate that INT and EXT disorders are partially driven by genetic differences, with heritability estimates ranging from 20-74% for EXT [9] and 32-37% for INT disorders [10, 11]. Large-scale genome-wide association studies (GWAS) have further revealed single-nucleotide polymorphism (SNP)-based heritability estimates, ranging from 5-22% for EXT behaviors [12, 13] and 11-20% for INT phenotypes [14]. These findings have enabled the generation of polygenic scores (PGS), which serve as quantitative indices of genome-wide liability for a given phenotype in independent samples at the individual level [15]. PGS for INT have shown modest predictive utility, explaining approximately 0.5-4.6% of INT phenotypes variance [16, 17], and PGS for EXT 10.5% of EXT phenotypes variance in adults most genetically similar to European ancestry [18].

Although INT and EXT disorders frequently co-occur and show moderate genetic correlations (rg≈ 0.44) [19], most genetic studies have analysed them independently. This approach has limited the application of models that consider shared liability for these disorders. Recent advances in statistical genetics, particularly genomic structural equation modelling (genomic SEM) [20], provide a powerful solution by enabling the modelling of latent genetic factors that capture both shared and trait-specific liability across correlated phenotypes. While genomic SEM has been used to investigate complex traits, few studies have leveraged this approach to identify pleiotropic genetic variants that could explain comorbidity between INT and EXT traits. This represents a significant gap, particularly given the mounting evidence of horizontal pleiotropy across INT and EXT genetic liabilities [21].

Although youth higher in genetic liability for INT and EXT disorders may be at heightened risk for experiencing either or both of these conditions, not all will develop these disorders. Decades of empirical and theoretical scholarship have highlighted the role of adverse childhood experiences and environments on INT and EXT psychopathology development [22]. For example, more frequent, chronic, and severe childhood adversity (e.g., abuse, neglect, poverty, caregiver mental illness, and family separation) is associated with an increased risk of developing INT, EXT, and comorbid INT-EXT disorders [23]. There is also evidence that environmental exposures may impact risk for INT and EXT disorders differently. For example, parental discord and parental depression have been linked to increased risk for INT disorders, and harsh parenting and physical punishment have been associated with heightened risk for EXT disorders specifically [24]. These findings highlight the importance of considering a range of environmental stressors that may exacerbate or attenuate the links between INT and EXT genetic liability and the manifestation of these conditions.

The current study aimed to address numerous limitations in the literature. First, we sought to identify the association between specific and common genetic liability for INT and EXT traits with INT and EXT disorder phenotypes in a sample of Mexican adolescents. Most GWAS to date have included predominantly individuals of European ancestry. Differences in allele frequencies, linkage disequilibrium, and environmental exposures observed across populations may influence the predictive utility of PGS derived from European ancestry to non-European ancestry samples [25]. Thus, determining the generalizability of genetic discoveries observed in European ancestry populations to admixed populations, such as Mexican youth, is sorely needed. Second, we examined whether adversity exposure moderated links between specific and shared genetic risk for INT and EXT disorders and the expression of these phenotypes. The distribution and impact of adversities in higher-income countries may differ substantially from those in low- and middle-income countries, such as Mexico. This difference potentially shapes distinct patterns of risk and resilience [26], underscoring the importance of examining the interplay between genes and environment in this population. Such work may inform interventions aimed at preventing or mitigating the emergence of INT and EXT phenotypes.

## Methods

### Participants and procedure

This study leveraged data from an epidemiological study (N=3,005) conducted in 2005 by the National Institute of Psychiatry in Mexico City and the Metropolitan area [27]. The sample includes adolescents who completed surveys regarding their mental health. Participants were 12-17 years old and were interviewed face-to-face in their homes by trained personnel using the computer-assisted World Mental Health Composite International Diagnostic Interview for Adolescents (WMH-CIDI-A) [27]. This structured interview generates psychiatric diagnoses based on the Diagnostic and Statistical Manual of Mental Disorders (4th ed.; 1994).

The analytic sample consisted of 1,130 adolescents who completed the diagnostic interview and had valid genetic data (Table 1). A comparison between the analytic sample and the rest of the cohort (i.e., those who either did not consent to provide mouthwash sample, had insufficient genetic material for genomic analysis, or whose data did not pass quality control) revealed no significant differences with respect to age (*t*(4126)= 0.68, *p*≥ 0.05) and sex (*χ***²**(1)= 2.26, *p*≥ 0.05), suggesting that the analytic sample is representative of the larger cohort.

**Table 1.** Sociodemographic, Clinical and Adversity Characteristics of the Sample.

|  |  |
| --- | --- |
|  | Total (N=1130) |
| Female | 620 (54.9%) |
| Male | 510 (45.1%) |
| Age (years) | 14.3±1.71 |
| Disorders |  |
| None | 554 (49.0%) |
| Internalizing only | 215 (19.0%) |
| Externalizing only | 214 (19.0%) |
| Comorbid Internalizing/Externalizing | 157 (13.0%) |
| Adversities |  |
| Neglect/Abuse | 222 (19.6%) |
| Parental loss | 373 (33.0%) |
| Parental psychopathology | 408 (36.1%) |
| Cumulative adversity (mean, SD) | 1.41 (1.30%) |

**Table 2.** Associations Between PGS and Internalizing Disorders, Externalizing Disorders, and Their Comorbidity.

|  | Internalizing |  | Externalizing |  | Comorbid<br>Internalizing/Externalizing |  |
| --- | --- | --- | --- | --- | --- | --- |
|  | OR [95%CI] | FDR | OR [95%CI] | FDR | OR [95%CI] | FDR |
| INT-SF | 0.97 [0.82-1.14] | 0.710 | 1.14 [0.96-1.36] | 0.371 | 1.05 [0.87-1.27] | 0.710 |
| EXT-SF | 1.02 [0.85-1.21] | 0.855 | 1.23 [1.03-1.47] | 0.070 | 1.09 [0.89-1.33] | 0.605 |
| INT-EXT-F | 1.18 [0.99-1.4] | 0.069 | **1.39 [1.16-1.67]** | **0.001** | **1.42 [1.16-1.75]** | **0.001** |

Notes. PGS= Polygenic Score INT-SF= internalizing-specific factor; EXT-SF= externalizing-specific factor, INT-EXT-F= shared internalizing–externalizing. All models were adjusted for age, sex, and genetic ancestry principal components. p-values were adjusted for multiple comparisons using the false discovery rate (FDR)< 0.05. Statistically significant interactions are presented in bold.

### Ethical Considerations

This study adhered to the ethical principles outlined in the Declaration of Helsinki [27] and was reviewed and approved by the Ethics and Scientific Committees of the National Institute of Psychiatry in Mexico City.

### Internalizing and Externalizing Disorder Assessment

Adolescents’ lifetime history of psychiatric disorders was assessed using the WMH-CIDI-A based on both adolescent self-reports and parent interviews. Diagnoses were determined according to DSM-IV criteria. A lifetime history of EXT disorders was defined as the presence of any of the following: Attention-Deficit/Hyperactivity Disorder (ADHD), Conduct Disorder, or Oppositional Defiant Disorder diagnosed based on adolescent’s self-reports and the parent interview, and Intermittent Explosive Disorder or Substance Use Disorders (SUD) based on adolescent reports only [27].

A lifetime history of INT disorders was based on adolescent self-reports, from which diagnoses of Major Depressive Disorder, Dysthymia, Panic Disorder, Generalized Anxiety Disorder, Social Phobia, Agoraphobia, and Separation Anxiety Disorder were derived [27].

Based on the presence or absence of INT and EXT disorders, participants were assigned to one of four mutually exclusive categories: 0= no diagnoses of these disorders; 1= INT disorders only; 2= EXT disorders only; and 3= presence of both INT and EXT disorders.

### Adversity Exposure

Childhood adversity was assessed through adolescent self-reports using the childhood section of the WMH-CIDI-A [28]. Adolescents reported experienced the following childhood adversities grouped into four categories based on previous work [27]: (1) neglect and abuse (neglect, physical abuse and sexual abuse); (2) loss (parental death, parental divorce and other separation from parents or caregivers); (3) parental psychopathology (mental disorders, substance use disorders, violence and criminal behavior); and (4) other adversities (family economic adversity and life-threatening childhood physical illness in the respondent). We examined three adversity factors (i.e., neglect and abuse, loss, and parental psychopathology), and cumulative adversity (including the other adversities category) as moderators of the association between polygenic liability for INT and EXT disorders. Cumulative adversity was categorized as None (0), Moderate (1–3), and High (≥4).

### Genomic Structural Equation Modelling

We used genomic SEM [20] to model the genetic factor structure underlying INT, EXT, and their comorbidity. The genetic variance-covariance matrix for a set of traits is estimated using linkage disequilibrium score regression (LDSC), which is then used to fit a user-specified model.

Publicly available GWAS summary statistics for 12 phenotypes were used: five internalizing traits (i.e., major depressive disorder diagnosis, anxiety disorder diagnosis, major depressive symptoms, generalized anxiety disorder symptoms, and neuroticism) [29–31] and seven externalizing traits (i.e., ADHD, risk tolerance, lifetime cannabis use, problematic alcohol use, smoking initiation, number of sexual partners, and age at first sexual intercourse) [18]. Where the original GWAS included the 23andMe cohort, summary statistics excluding it were used in the current study. Information regarding these GWAS can be found in Supplemental Table S1.

The genetic covariance matrix for the 12 traits was estimated using multivariable LDSC (*ldsc* function in genomic SEM R package). SNP-based heritability estimates were transformed to the liability scale for case-control traits to ensure comparability across continuous and dichotomous phenotypes before model estimation. We employed the approach recommended by Grotzinger et al. [32], where sample prevalence is set to 0.5 and the effective sample size of the trait (instead of the total sample size) is specified (see Supplemental Table S1 for effective N and population prevalences). We evaluated four models: (1) a common factor model, which assumes a single latent dimension of genetic liability; (2) a two-correlated factors model, which represent distinct, but correlated, INT and EXT dimensions; (3) a hierarchical model, with a higher-order general factor and lower-order INT and EXT factors; and (4) a bifactor model, with a general factor and orthogonal INT- and EXT-specific factors.

Model fit was evaluated using the Comparative Fit Index (CFI), Standardized Root Mean Square Residual (SRMR), and Akaike Information Criterion (AIC). Acceptable model fit was defined as CFI≥ 0.90 and SRMR≤ 0.08, consistent with established guidelines [20]. Following model selection, we conducted a multivariate GWAS using genomic SEM. Analysis included 3,159,283 SNPs that are common to all GWAS and the 1000 Genomes Phase 3 reference panel [33], as well as with minor allele frequency (MAF)> 0.01. Genome-wide significance was set at *p*< 5×10⁻⁸. PLINK 1.9 [34] was used for LD clumping (*r²*< 0.10).

### PGS Construction

PGS were computed using SBayesR [35]. This Bayesian multiple regression method accounts for LD structure and improves the accuracy of polygenic prediction. GWAS summary statistics from the multivariate GWAS of the latent factors were used to derive the PGS. Quality control procedures included the exclusion of SNPs with imputation quality *r²*< 0.6, call rates below 0.90, MAF< 0.01, non-autosomal SNPs, and strand-ambiguous variants. The resulting scores were standardized as z-scores and used as predictors in all subsequent regression analyses.

To control for population stratification, we conducted principal component analysis on genotype data using PLINK 2.0 (v2.00a3.3) [36]. Individuals with greater than 5% missing genotypes were excluded, and variants were filtered for missingness (>5%) and MAF (<1%). LD pruning was applied to retain a subset of independent SNPs. The top ten ancestry principal components (PCs) were extracted and included as covariates in all models to adjust for residual population structure [37].

### Statistical Analyses

We used multinomial logistic regression to examine associations between PGS derived from the best-fitting genomic SEM and corresponding phenotypic disorders. All models included sex, age, and the first ten genetic ancestry PCs. Model fit was assessed using log-likelihood ratio tests. To quantify model-level explanatory power, we computed McFadden’s pseudo-R², a likelihood-based measure that reflects the proportion of variance in diagnostic outcomes explained by each model relative to a null (intercept-only) model [38]. Pseudo R² values were used to compare explanatory strength across models, particularly when evaluating the incremental value of adversity and gene-by-environment (G×E) interactions.

We first examined the main effects of INT and EXT PGS on the individual and joint expression of these phenotypes. We then extended these models to include the main effects of three adversity domains—neglect/abuse, parental loss, and parental psychopathology—to evaluate their independent associations with psychiatric diagnoses. To assess moderation, we first included interaction terms between PGS and each adversity domain, and then between PGS and the total number of adversities in an independent model. Following best practice recommendations for modeling G×E effects [36], in all GxE interaction models, sex, age, and genetic ancestry PCs were included. All interaction models also included cross-product terms between adversity variables and covariates (adversity × sex) and PGS and covariates (PGS x sex) to prevent spurious interaction estimates due to omitted variable bias.

Statistical significance was set at *p*< 0.05. To adjust for multiple comparisons, the Benjamini-Hochberg false discovery rate (FDR) procedure [39] was applied separately within each predictor term, allowing for targeted control of Type I error. All analyses were conducted in Rv4.3.0 [40], using the tidyverse, nnet, ggplot2, and poLCA packages for data processing, modelling, and visualization.

## Results

### Genetic Factor Structure of Internalizing and Externalizing Traits

Among the tested Genomic SEM models, the bifactor model provided the best fit to the data (AIC= 750.978, CFI= 0.973, SRMR= 0.052). Full model fit indices for all tested models are provided in Supplemental Table S2, and standardized factor loadings for the bifactor model are detailed in Supplemental Table S3. The bifactor model (Supplemental Figure S4) partitioned genetic covariance into a common EXT-INT factor (INT-EXT-F; capturing shared genetic variance across INT and EXT traits), an INT-specific factor (INT-SF; accounting for genetic influences unique to INT traits), and an EXT-specific factor (EXT-SF; representing genetic contributions unique to EXT traits). Multivariate GWAS on these latent factors identified 224 loci for INT-EXT-F, 151 loci for INT-SF, and 42 loci for EXT-SF.

### Associations of PGS for Internalizing and Externalizing Traits and Internalizing and Externalizing Disorders

The main effects PGS model explained a small proportion of variance (pseudo R²= 0.0385) in the INT and EXT disorders (Supplemental Table S5). Using multinomial logistic regression, we examined the associations between PGS (i.e., INT-EXT-F, INT-SF, EXT-SF) and phenotypic INT and EXT disorders, as well as their co-occurrence. Greater INT-EXT-F PGS was significantly associated with increased odds of EXT only disorders (OR= 1.39; FDR< 0.001) and comorbid INT-EXT disorders (OR= 1.42; FDR< 0.001) (Figure S6). No other main effects withstood FDR correction. Full details regarding regression coefficients and ORs for all PGS-outcome associations are reported in Table 3.

**Table 3.** Gene-by-Environment Interactions Between PGS and Adversity in Predicting Phenotypic Internalizing Disorders, Externalizing Disorders, and Their Comorbidity.

|  | Internalizing |  | Externalizing |  | Comorbid Internalizing/<br>Externalizing |  |
| --- | --- | --- | --- | --- | --- | --- |
|  | OR [95%CI] | FDR | OR [95%CI] | FDR | OR [95%CI] | FDR |
| INT-SF x<br>neglect/abuse | 0.84 [0.5-1.41] | 0.766 | 0.78 [0.491-1.25] | 0.766 | 1.06 [0.66-1.72] | 0.806 |
| INT-SF x<br>parental loss | 1.32 [0.87-1.98] | 0.285 | 1.41 [0.93-2.13] | 0.285 | 1.1 [0.7-1.74] | 0.676 |
| INT-SF x<br>parental<br>psychopathology | 0.85 [0.63-1.14] | 0.389 | 0.88 [0.66-1.18] | 0.389 | 0.76 [0.56-1.03] | 0.238 |
| INT-SF x<br>cumulative adversity | 0.99 [0.84-1.17] | 0.948 | 0.99 [0.841-1.15] | 0.948 | 1.01 [0.85-1.19] | 0.948 |
| EXT-SF x<br>neglect/abuse | 1.58 [0.95-2.63] | 0.118 | 1.75 [1.06-2.87] | 0.082 | 1.25 [0.75-2.1] | 0.392 |
| EXT-SF x<br>parental loss | **1.76 [1.17-2.64]** | **0.017** | 1.16 [0.77-1.73] | 0.481 | **1.79 [1.14-2.81]** | **0.017** |
| EXT-SF x<br>parental<br>psychopathology | 0.79 [0.56–1.1] | 0.265 | 0.96 [0.7-1.32] | 0.814 | 1.27 [0.9-1.79] | 0.265 |
| EXT-SF x<br>cumulative adversity | **1.18 [1.00-1.4]** | **0.049** | **1.2 [1.01-1.42]** | **0.049** | **1.24 [1.04-1.48]** | **0.042** |
| INT-EXT-F x<br>neglect/abuse | 1.48 [0.88-2.48] | 0.417 | 1.06 [0.65-1.74] | 0.814 | 0.89 [0.53-1.49] | 0.814 |
| INT-EXT-F x<br>parental loss | 1.63 [1.09-2.44] | 0.050 | 1.1 [0.73-1.64] | 0.865 | 1.04 [0.66-1.64] | 0.865 |
| INT-EXT-F x<br>parental<br>psychopathology | 0.89 [0.64-1.23] | 0.779 | 0.9 [0.65-1.25] | 0.779 | 1.02 [0.73-1.44] | 0.896 |
| INT-EXT-F x<br>cumulative adversity | **1.27 [1.07-1.5]** | **0.021** | 1.06 [0.89-1.27] | 0.735 | 0.99 [0.83-1.19] | 0.934 |

Note. Each row reports the interaction term between a specific PGS—the internalizing-specific factor (INT-SF), externalizing-specific factor (EXT-SF), or shared internalizing–externalizing factor (INT-EXT-F) and an adversity exposure. P-values were adjusted for multiple comparisons using the false discovery rate (FDR) < 0.05. Statistically significant interactions are presented in bold.

### PGS x Adversity Interactions in Predicting Internalizing and Externalizing Disorders

Including interaction terms increased the variance accounted for in the outcomes, with the full interaction model achieving the highest explanatory power (pseudo R²= 0.1625), followed by the cumulative adversity model (pseudo R²= 0.1275). Numerous interactions involving adversity and INT-SF and EXT-SF PGS, as well as these phenotypes, were noted. Parental loss moderated the association between EXT-SF PGS and both INT only (OR= 1.76; FDR< 0.05) and INT-EXT comorbidity (OR= 1.79; FDR< 0.05), indicating that individuals with higher EXT-SF PGS had an increased odds of INT disorders only and co-occurring INT-EXT disorders when exposed to parental loss (Figure 1). Furthermore, cumulative adversity moderated associations of EXT-SF PGS with psychiatric outcomes, such that higher EXT-SF PGS was associated with an increased odds of INT disorders only (OR= 1.18; FDR< 0.05), EXT disorders only (OR= 1.20; FDR< 0.05), and comorbid INT-EXT disorders (OR= 1.24; FDR< 0.05) (Figure 2). The cumulative number of adversities also interacted with the INT-EXT-F PGS in predicting risk for INT disorders only (OR= 1.27; FDR< 0.05). As shown in Figure 3, adolescents with greater genetic liability on the common INT-EXT factor were at increased risk for experiencing INT disorders, but only when exposed to a greater number of adversities. Full results from these analyses are presented in Table 3 and Supplemental Table S5.

**Figure 1.**
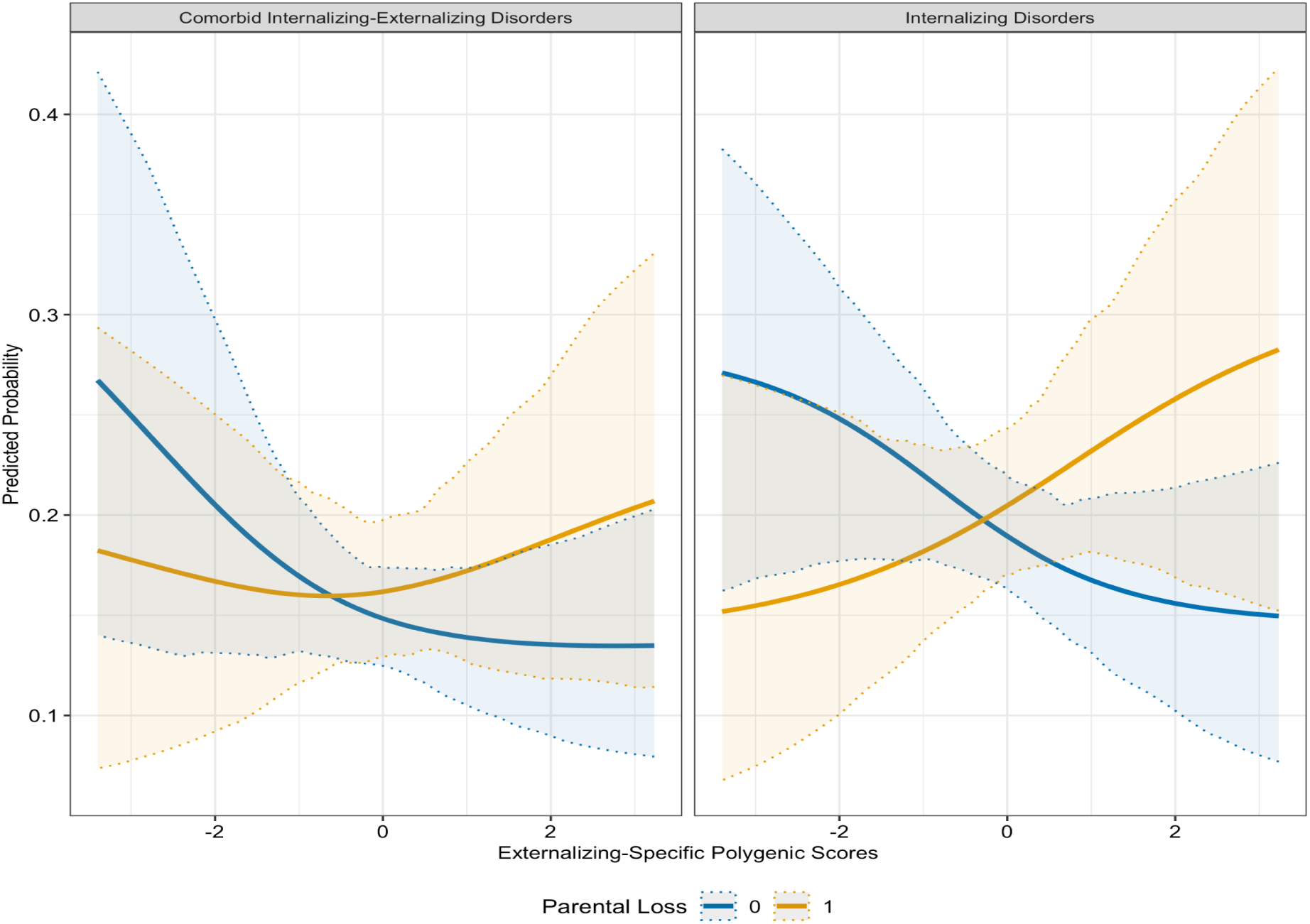
Interaction Between Externalizing-Specific Polygenic Scores and Parental Loss in Predicting Internalizing Disorders and Internalizing/Externalizing Comorbidity

**Figure 2.**
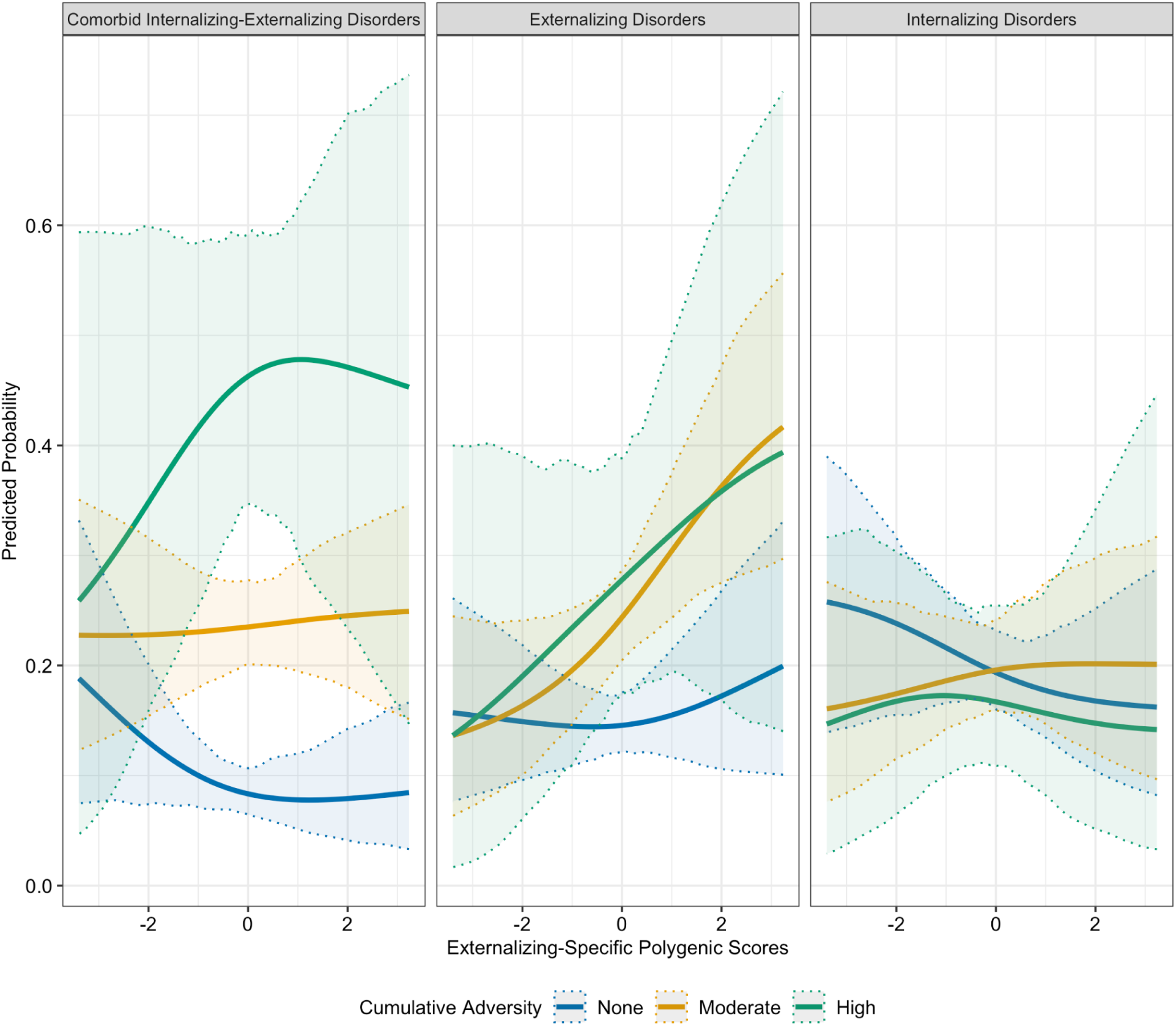
Cumulative Adversity Moderates Genetic Effects on Risk for Individual and Comorbid Internalizing and Externalizing Disorders

**Figure 3.**
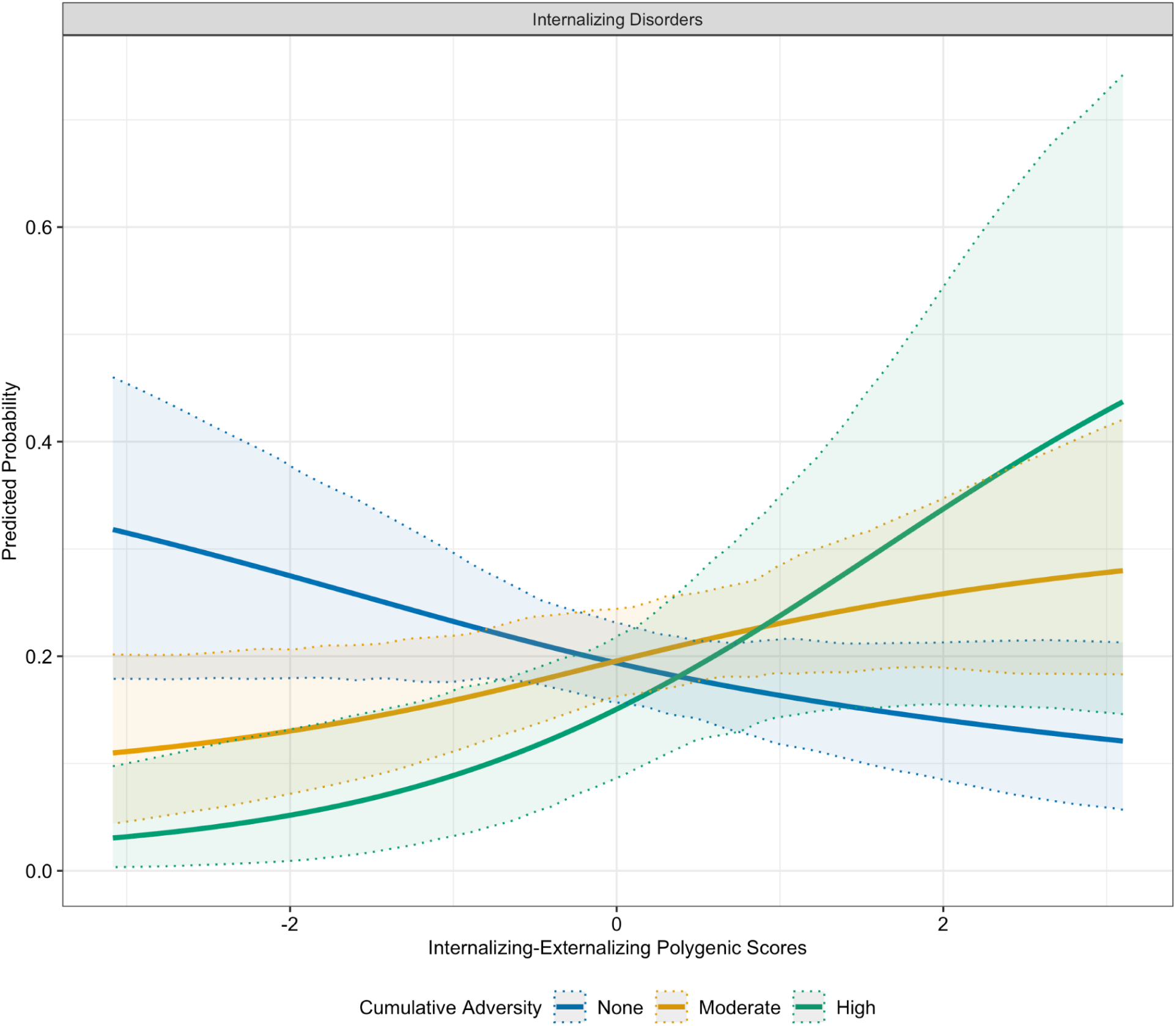
Interaction Between Shared Genetic Liability for Internalizing and Externalizing Disorders and Cumulative Adversity in Predicting Internalizing Disorders

## Discussion

The present study examined the relations between specific and shared genetic risk for INT and EXT traits on the expression of corresponding disorders, and explored the moderating role of adversity exposure on these links in a population-based sample of Mexican adolescents, building on the limited work in this area. Genomic SEM suggested that a bifactor model best depicts the genetic structure underlying INT and EXT traits, which contains a general latent factor (INT-EXT-F), alongside orthogonal specific factors for internalizing (INT-SF) and externalizing (EXT-SF) dimensions. The identified bifactor model from genomic SEM supports a dimensional framework of psychopathology, suggesting that common risk factors underlie INT and EXT phenotypes [20, 41], helping to clarify the genetic architecture underlying these conditions.

PGS derived from genomic SEM revealed that INT-EXT-F was robustly associated with EXT-only and comorbid INT-EXT diagnoses, but not with INT-only diagnosis. The link between PGS for INT-EXT-F and the phenotypic manifestations of EXT-only and comorbid INT-EXT disorders suggests that much of the genetic risk underlying phenotypic EXT and comorbid INT-EXT disorders is conveyed through a common predisposition to INT and EXT disorders.

Contrary to expectations, we did not observe significant main effects of the INT-SF PGS on INT-only disorders or the EXT-SF PGS on EXT-only disorders, which contrasts with findings from prior analyses in this sample [42]. This can likely be explained by differences in statistical power between the common and specific PGS factors. Our multivariate GWAS approach leverages the shared genetic architecture across multiple phenotypes, which boosts statistical power to detect variants implicated in the common liability factor (INT-EXT-F) [18]. Conversely, the specific factors (INT-SF and EXT-SF) are derived from the residual genetic variance after accounting for shared influences, and PGS based on these residuals tend to explain less variance in their target traits. This is consistent with prior research showing that PGS based on common liability factors often outperform those based on trait-specific residuals in prediction [42].

Another potential explanation, particularly for the null finding for the INT-SF, is the developmental dynamism of genetic influences on INT psychopathology. The GWAS summary statistics used to create the PGS were derived from adult cohorts. While common genetic factors for EXT tend to show stability from a young age [41], genetic influences on INT problems show less continuity across development [43]. For instance, twin studies indicate that while genetics account for nearly 75% of the variance in anxiety and depression in middle childhood, this same genetic factor explains only 12% of the variance in young adulthood [43]. Therefore, the adult-derived INT-SF PGS may not adequately capture the specific genetic risk for internalizing disorders during adolescence, underscoring the critical need for gene-identification efforts in younger cohorts.

We also found that parental loss moderated the association between EXT-SF PGS and phenotypic INT-only and comorbid INT-EXT disorders. These findings illustrate the concept of multifinality, or that the same environmental exposure (e.g., parental loss) can lead to divergent outcomes [44]. The absence of a caregiver, through either divorce or death, is typically a major life stressor for parents and children alike that may have detrimental effects on dyadic relationships and the family system as a whole [45]. The loss of a caregiving partner can negatively impact parents’ mental health (e.g., increased risk for depression) or interfere with the provision of positive parenting practices (e.g., warmth, responsiveness, scaffolding of emotion regulation skills). Such experiences may be particularly detrimental among high genetic liability youth for EXT symptoms who tend to have more challenges with self-regulation. Indeed, it appears that children with higher EXT-SF genetic liability who face the loss of a caregiver may experience greater INT, or comorbid INT-EXT disorders. A plausible hypothesis is that some children with high EXT-SF genetic liability may act out due to difficulties in navigating disruptions in the family unit and experience co-occurring INT-EXT symptoms, whereas some social and familial norms may discourage children from expressing EXT behaviors, leading them to internalize feelings of distress when exposed to parental loss [46]. Future research is needed to understand the factors associated with differential adaptation among children with higher EXT-SF genetic liability in the context of parental loss.

Numerous interactions were observed between cumulative adversity and EXT-SF PGS with INT-only, EXT-only disorders, and their co-occurrence. Cumulative adversity also moderated associations between INT-EXT-F PGS and phenotypic INT-only disorders. In general, we observed a dose-dependent pattern, whereby the probability of INT-only, EXT-only, and comorbid outcomes increased progressively with both higher genetic liability and greater exposure to adversity. These results broadly align with cumulative stress models of psychiatric risk [47] and suggest that genetic vulnerabilities may become more penetrant in the context of increasing environmental burden. Individuals showing higher EXT-SF PGS might be more vulnerable to experience deficits in self-regulation and a greater number of adverse childhood experiences, which could interfere with brain development, specifically neural areas underpinning cognitive control and affective regulation. These adverse experiences can increase risk for INT and EXT disorders and their comorbidity. It is also possible that greater adversity exposure may interfere with the completion of age-expected developmental tasks in academic or interpersonal domains; this may be especially true for youth with a higher genetic liability for EXT-SF or INT-EXT-F who are predisposed to experiencing affective and behavioral dysregulation, thereby increasing their risk for INT and EXT behaviors.

Several limitations warrant mention. First, GWAS summary statistics used to estimate PGS were drawn from samples most genetically similar to European ancestry populations, which may have limited the predictive utility of the PGS in our target population (Mexican adolescents). Future genetic studies efforts should consider using diverse ancestry cohorts. Second, the study design was cross-sectional, precluding us from drawing causal inferences. Future longitudinal studies are essential to examine the impact of adversity and genetic liability for INT and EXT on developmental trajectories of these phenotypes. Lastly, GWAS summary statistics used to derive the PGS were from adult populations. To our knowledge, no large-scale GWAS has been conducted on INT and EXT conditions in adolescent populations. As mentioned previously, genetic loci underpinning INT and EXT traits identified among adults may lose statistical power in adolescent samples, attenuating the associations observed here. An important avenue for future research is the inclusion of younger individuals, which may enhance the predictive power of the INT and EXT PGS in younger samples.

In conclusion, our results provide insight into the genetic etiology of INT disorders, EXT disorders, and their comorbidity. Common liability INT-EXT genetic factors underlie the comorbidity of INT and EXT disorders, as well as EXT disorders specifically. We also found evidence that EXT genetic liability contributes to INT and INT-EXT comorbidity among adolescents who are exposed to parental loss or multiple adversities. Although the clinical applications of our findings are currently limited, incorporating PGS into psychiatric settings for risk assessment, alongside the consideration of traditional clinical risk factors, has several promising applications [48]. Future research is needed to determine whether early screening and interventions aimed at attenuating risk for internalizing symptoms–or comorbidity of these symptoms with externalizing behaviors–may benefit youth higher in genetic risk for EXT and have experienced parental loss or multiple life adversities. Such work may inform more targeted risk stratification and assessment.

## Data Availability

Summary-level data and code used as part of this project can be obtained from the authors upon reasonable request. Sharing of individual data is restricted due to ethical requirements and to protect the participant's privacy. Enquiries should be directed to the authors.

